# Multimodal Ageing Biomarkers and Plasma Proteomic Signatures Associated with All-Cause Mortality

**DOI:** 10.64898/2026.03.09.26347914

**Authors:** Maira Pyrgioti, Ines Mesa Eguiagaray, Paul Redmond, Janie Corley, Mark E. Bastin, Maria Valdés Hernández, Tom C. Russ, Joanna M. Wardlaw, Eilis Hannon, Ian J. Deary, Keenan A. Walker, Elliot M. Tucker-Drob, Simon R. Cox, Riccardo E. Marioni, Sarah E. Harris

## Abstract

Ageing biomarkers can predict mortality risk beyond chronological age. Recently, plasma proteins were used to estimate the biological ages of eleven human organs, including the brain, heart, liver, kidneys, and pancreas. Accelerated organ ageing is linked to higher all-cause mortality; however, systematic benchmarking against established ageing biomarkers is lacking. Here, we pursued two complementary aims. First, we benchmarked proteomic organ ages against multimodal ageing biomarkers for all-cause mortality (444 deaths; ≤17-year follow-up) using Cox regression in 861 Lothian Birth Cohort 1936 (LBC1936) participants. Ageing biomarkers included epigenetic age (GrimAge2), telomere length, neuroimaging, general cognitive function (*g*), and physical function (grip strength, walk time, and respiratory function). Among proteomic organ ageing biomarkers, accelerated liver (HR_perSD_ [95%CI] = 1.43 [1.30–1.58]), immune (1.42 [1.29–1.57]), and heart (1.38 [1.25–1.53]) ageing were most strongly associated with higher mortality risk. However, GrimAge2 acceleration, total brain volume (TBV), grey matter volume, respiratory function, and *g* exhibited higher hazard estimates (HR_perSD_ = 1.44–1.62) than organ ageing biomarkers. In a Cox model including all biomarkers, only TBV, white matter hyperintensity volume, *g*, and walk time associated with mortality. Second, survival analyses of SomaScan 11K plasma proteins identified 202 proteins associated with mortality and enriched for the liver and immune-related biological processes, with the strongest effects observed for GDF15 (HR_perSD_ [95%CI] = 1.53 [1.37–1.72]), CST3 (1.48 [1.29–1.69]), and COL18A1 (1.47 [1.30–1.68]). These findings provide a systematic, cross-modal benchmarking of proteomic organ ages against established ageing biomarkers and highlight plasma proteomic signatures of mortality.

**GRAPHICAL ABSTRACT:** 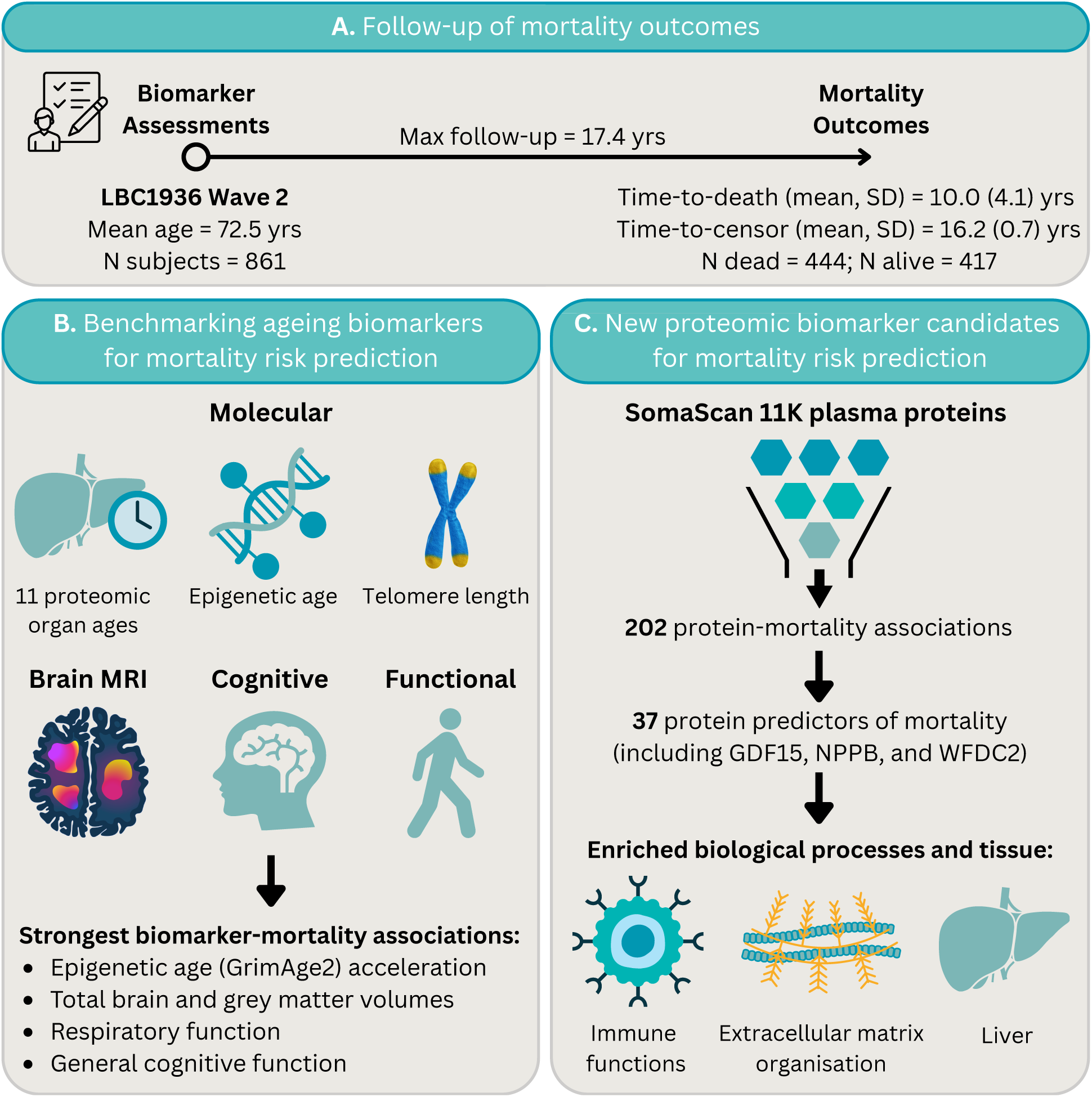

## 1 Introduction

There is a pressing clinical need to develop biomarkers of ageing—measurable biological parameters that capture age-related changes and outperform chronological age in predicting age-related outcomes (Aging Biomarker Consortium, 2023). Such biomarkers could improve mortality risk stratification, serve as surrogate endpoints in longevity trials, and guide personalised health interventions. Multiple biomarker modalities have emerged, including DNA-based, epigenetic, transcriptomic, proteomic, metabolomic, functional, and neuroimaging markers, which enhance risk prediction beyond chronological age; however, each modality has notable limitations (Argentieri et al., 2024; Cooper et al., 2010; Ho et al., 2023; Ibáñez de Opakua et al., 2025; Jiang et al., 2017; Jylhävä et al., 2017; Kuo et al., 2024; Li et al., 2020; Marioni et al., 2015; Mavrommatis et al., 2025; Tanaka et al., 2020; Wang et al., 2018).

Telomere length—a marker of replicative senescence—shows only a weak inverse correlation with chronological age and inconsistent associations with mortality across studies, corresponding to an average 9% higher mortality risk per standard deviation decrease (Levy et al., 1992; Wang et al., 2018; Ye et al., 2023). Functional biomarkers are easy to measure and consistently associate with mortality, yet they provide no mechanistic insight into ageing biology and typically show modest effect sizes. For instance, lower grip strength and slower walking speed are associated with 27% and 14% higher mortality risk per standard deviation or 0.1 m/s decrease, respectively (Gale et al., 2007; Studenski et al., 2011). In contrast, neuroimaging markers capture brain structural changes and demonstrate stronger associations with mortality, with lower total brain volume and greater white matter hyperintensity volume corresponding to 52% and 32% higher risk per standard deviation change, respectively (Van Elderen et al., 2016; Ghaznawi et al., 2021). While informative, neuroimaging measures are resource-intensive to generate and do not directly reflect molecular ageing processes, highlighting the need for biomarkers that are both mechanistically insightful and broadly predictive of ageing and survival.

Advances in high-throughput omics technologies have brought a new generation of ageing biomarkers to the fore. DNA methylation patterns change systematically with age and contribute to age-related disease (Vaidya et al., 2025). Epigenetic clocks, which use machine learning models trained on DNA methylation data to predict chronological or biological age, show robust associations with mortality (Bocklandt et al., 2011; Hannum et al., 2013; Horvath, 2013; Horvath & Raj, 2018; Lu et al., 2022), with up to a 54% increased risk per standard deviation of age acceleration (Mavrommatis et al., 2025). However, they typically provide a single, systemic measure of ageing. Transcriptomic studies in animals have revealed conserved ageing signatures across organs, alongside organ-specific differences in temporal trajectories (Schaum et al., 2020). Despite the potential of organ ageing measures to improve understanding of organ-specific disease and mortality risk at the individual level, organ-level heterogeneity in human ageing has only recently begun to be characterised. Oh et al. introduced an approach that leverages organ-enriched plasma proteomic signatures to estimate the biological ages of eleven major organs, including the brain, heart, immune system, kidneys, liver, adipose tissue, and pancreas (Oh et al., 2023). Most proteomic organ age measures were significantly associated with all-cause mortality in the LonGenity cohort, with each standard deviation increase in organ age corresponding to a 17–53% higher risk of death (Oh et al., 2023). However, their comparative performance for mortality prediction against other ageing biomarkers within the same population remains unknown. This comparison is critical to determine whether organ clocks provide incremental value for mortality risk prediction beyond existing biomarkers and to prioritise the most informative biomarkers for clinical risk stratification.

Here, we pursue two complementary aims. First, we systematically benchmark the eleven proteomic organ ages against other molecular, physical function, cognitive, and neuroimaging biomarkers of ageing for predicting all-cause mortality in 861 Lothian Birth Cohort 1936 (LBC1936) participants (n=444 deaths; ≤17-year follow-up), using Cox models adjusted for chronological age, sex and modifiable risk factors. This approach can elucidate the multi-systemic contributions to mortality. Second, we conduct survival analyses of 8,699 unique plasma proteins measured with the SomaScan 11K assay to identify novel proteomic candidates associated with longevity, and perform enrichment analyses to highlight overrepresented biological processes and tissues. We further apply elastic net-penalised Cox regression to derive a concise set of proteins associated with mortality. This integrated, multimodal benchmarking approach reveals the most informative ageing biomarkers, proteins, and biological pathways underlying mortality risk.

## 2 Materials and Methods

### 2.1 Study Population

The LBC1936 is a longitudinal study of ageing trajectories in individuals born in 1936 who participated in the Scottish Mental Survey at age 11 and were residing in Scotland at the time of recruitment. Participants underwent repeated, comprehensive assessments of cognitive function, brain imaging, physical health, and fluid biomarkers approximately every three years, between the ages of ∼70 and ∼88 years. SomaScan 11K (v5.0) plasma proteomics and brain imaging were introduced at wave 2 (mean age = 73 years). Mortality status was ascertained via linkage to the National Health Service Central Register (NHSCR), managed by the National Records of Scotland (NRS), which provides regular updates. The mortality outcomes used in the present study were last updated on 9 September 2025. Detailed cohort characterisation and methodologies have been described previously (Deary, Pattie, & Starr, 2012; Taylor, Pattie, & Deary, 2018). All baseline biomarker measurements for the present study were obtained from wave 2 and are summarized in Table 1. Many of these non-organ-age biomarkers have previously been associated with mortality in this cohort, albeit with shorter follow-up periods and lower mortality incidence (Cole et al., 2018; Corley et al., 2019; Harris et al., 2016; Hillary et al., 2021; McLachlan et al., 2020).

**TABLE 1.** Baseline characteristics of LBC1936 (wave 2)

| Variables | Mean (SD) | N Subjects |
| --- | --- | --- |
| Demographic factors |  |  |
| Chronological age (years) | 72.5 (0.7) | 861 |
| Male/female | N/A | 445/416 |
| Time-to-death (years) | 10.0 (4.1) | 444 |
| Time-to-censor (years) | 16.2 (0.7) | 417 |
| Modifiable risk factors |  |  |
| Smoking pack years | 15.4 (24.7) | 850 |
| Alcohol units per week | 11.7 (14.4) | 769 |
| BMI (kg/m<sup>2</sup>) | 27.9 (4.5) | 861 |
| Physical function measures |  |  |
| Six meter walk time (s) | 4.4 (1.3) | 855 |
| Grip strength (kg) | 28.5 (9.4) | 818 |
| FEV (L) | 2.3 (0.7) | 851 |
| FVC (L) | 3.1 (0.9) | 851 |
| FER (%) | 77.4 (11.7) | 851 |
| Brain MRI volume measures |  |  |
| TBV/ICV | 0.7 (0.02) | 656 |
| GMV/ICV | 0.3 (0.02) | 656 |
| WMHV/ICV | 0.008 (0.009) | 666 |
| Molecular measures |  |  |
| Leukocyte telomere length (bp) | 3969.7 (738.1) | 839 |

Notes: BMI = Body Mass Index, FEV = Forced Expiratory Volume in 1 second; FVC = Forced Vital Capacity; FER = Forced Expiratory Ratio; MRI = Magnetic Resonance Imaging; TBV/ICV = Total Brain Volume to Intracranial Volume ratio; GMV/ICV = Grey Matter Volume to Intracranial Volume ratio; WMHV/ICV = White Matter Hyperintensity Volume to Intracranial Volume Ratio.

### 2.2 Plasma proteomics

Plasma was extracted from blood collected in citrate tubes at waves 2 (age ∼73 years), 3 (age ∼76 years), 5 (age ∼82 years), and 6 (age ∼86 years). Protein quantification was performed using the SomaScan 11K (v5.0) platform, a high-throughput proteomic assay that employs slow off-rate modified DNA aptamers (SOMAmers) and a DNA microarray readout to measure 11,083 protein targets in relative fluorescence units (RFU) from 55 µL of plasma (Candia et al., 2024; Kirsher et al., 2025). Standard normalisation procedures, including hybridisation control normalisation, median signal normalisation on calibrators, plate-scale normalisation, interplate calibration, and median signal normalisation on all sample types, were performed by SomaLogic as previously described (Candia et al., 2025). Additional quality control (QC) procedures were applied to exclude non-human and non-protein aptamers, aptamers with a coefficient of variation (CV) greater than 50%, low-variance aptamers, aptamers with a median RFU above 80,000, and those with signal-to-noise ratios below 1. Principal component analysis revealed differences between waves, particularly in wave 2. Consequently, 963 aptamers exhibiting low correlation (<0.3) and high absolute mean difference (>0.5) between wave 2 and the other waves were removed. Cross-referencing these aptamers with SomaLogic pre-analytical variation (PAV) tables indicated that over 75% displayed moderate-to-high PAV effects. Following QC, 9,703 aptamers, corresponding to 8,699 unique UniProt IDs, were retained for LBC1936 wave 2 (N = 789). Protein expression values were log10-transformed and standardized (mean = 0, SD = 1) prior to downstream analyses.

### 2.3 Proteomic organ age gaps

Proteomic organ ages were derived in LBC1936 wave 2 using the organ-specific age predictors previously established by Oh et al. (2023). The organs comprised adipose tissue, artery, brain, heart, immune system, intestine, kidney, liver, lung, muscle, and pancreas. Organ-enriched transcripts were defined as those exhibiting at least four-fold higher expression in a given organ relative to all others, based on bulk tissue RNA-sequencing data from the Genotype-Tissue Expression (GTEx) Project. Tissues corresponding to the same organ were aggregated such that a gene’s expression level for each organ was defined by the maximum expression observed among its constituent subtissues (e.g., all GTEx brain regions were treated as subtissues of the brain organ). The immune organ was characterised using gene expression data from blood and spleen. These organ-enriched transcripts were subsequently mapped to 4,979 plasma proteins measured by the SomaScan assay, and 893 organ-enriched proteins were identified. After additional QC to remove proteins with a high coefficient of variation or a low correlation between SomaScan assay versions v.4 (5k panel) and v.4.1 (7k panel), 4,778 proteins were retained, corresponding to 856 organ-enriched proteins. Organ-specific age predictors were trained in 1,398 cognitively normal participants from the Knight Alzheimer Disease Research Center (Knight-ADRC) cohort. For each organ, bootstrap-aggregated LASSO regression models (500 bootstrap iterations per organ) were fitted with standardised (mean = 0, SD = 1), log10-transformed organ-enriched protein levels as predictors, chronological age as the outcome, and sex included as a covariate. For LBC1936 wave 2, log10-transformed SomaScan 11K protein measures were standardised (mean = 0, SD = 1) and aligned to the protein coefficients published by Oh et al. (2023). For each participant, predicted organ age was calculated by applying the 500 bootstrap sets of protein and sex coefficients and averaging the resulting predicted ages (bagging) to obtain a single age estimate per organ. Organ age gaps were defined as standardised (mean = 0, SD = 1) residuals from linear regression models of predicted organ age on chronological age, fitted separately for each organ. Positive age gaps indicate relatively accelerated organ ageing and negative values indicate relatively slower organ ageing compared with peers of the same chronological age.

### 2.4 GrimAge2 acceleration

DNA methylation (DNAm) profiling was performed on whole-blood LBC1936 wave 2 samples using the Illumina HumanMethylation450K BeadChip. Detailed protocols for quantification and QC of DNAm data in the LBC1936 have been reported previously (Zhang et al., 2018). DNAm GrimAge2 was estimated for LBC1936 samples using Horvath’s online calculator (https://dnamage.clockfoundation.org/). GrimAge2 was developed via a two-stage process (Lu et al., 2022). First, DNAm-based surrogates were established for smoking pack-years and nine plasma proteins: adrenomedullin (ADM), beta-2 microglobulin (B2M), cystatin C (CST3), growth differentiation factor 15 (GDF15), leptin (LEP), plasminogen activator inhibitor-1 (PAI-1), tissue inhibitor of metalloproteinases 1 (TIMP1), log-transformed high-sensitivity C-reactive protein (CRP), and log-transformed hemoglobin A1C (HbA1c). Second, a Cox regression model with elastic net regularisation was used to predict all-cause mortality, with time-to-death as the outcome and the nine DNAm protein surrogates, DNAm smoking pack-years, chronological age, and sex as predictors. The resulting uncalibrated DNAm GrimAge2 estimate was linearly transformed to units of years to serve as a physiological age estimator. GrimAge2 acceleration was calculated as the standardised residuals from a linear model regressing GrimAge2 on chronological age, with positive values indicating accelerated ageing and negative values indicating decelerated ageing.

### 2.5 Telomere length

DNA was extracted from peripheral blood leukocytes, and telomere length was measured by quantitative real-time polymerase chain reaction (qPCR) using an Applied Biosystems 7900HT Fast Real-Time PCR system (Pleasanton, CA, USA) (Martin-Ruiz et al., 2004). Four internal control DNA samples were run within each plate to calculate absolute telomere lengths and to account for inter-plate variation. These controls consisted of cell lines with known telomere lengths of 6.9 kb, 4.03 kb, 2.0 kb, and 1.32 kb. The relative telomere-to-glyceraldehyde 3-phosphate dehydrogenase (T/S) ratios of the controls were used to generate a standard regression line, which was then applied to convert the relative telomere length measurements of study samples into absolute telomere lengths. This methodology has been described in detail previously (Harris et al., 2016).

### 2.6 Physical function

Methods for measuring physical function biomarkers have been described previously (Deary et al., 2007; Okely & Deary, 2020). Walk time was assessed as the time (in seconds) taken to walk 6 metres along a corridor, measured with a stopwatch, with longer times indicating slower walking (Okely & Deary, 2020). Grip strength was measured three times in each hand using a North Coast Hydraulic Hand Dynamometer (JAMAR) (Deary et al., 2007; Okely & Deary, 2020), and the highest score from the three attempts with the dominant hand was used. Lung function was assessed using a Micro Medical Spirometer and included forced expiratory volume (FEV, L), defined as the volume of air expelled during the first second of a forceful exhalation; forced vital capacity (FVC, L), the total volume of air expelled during a forceful exhalation; and the forced expiratory ratio (FER, %), representing the percentage of air exhaled in the first second relative to the total exhaled volume (Deary et al., 2007; Okely & Deary, 2020). Participants were given three attempts on the spirometer, and the highest scoring attempt was used.

### 2.7 General cognitive function (*g*)

LBC1936 wave 2 participants completed 13 cognitive tests spanning four cognitive domains, as described in detail elsewhere (Deary et al., 2007; Moodie et al., 2026; Ritchie et al., 2016; Tucker-Drob et al., 2014). The domains and corresponding tests were as follows:

1. **Visuospatial Skills** – Block Design (Wechsler, 1997a), Matrix Reasoning (Wechsler, 1997a), and Spatial Span (Wechsler, 1997b).
2. **Processing Speed** – Symbol Search (Wechsler, 1997a), Digit-Symbol Substitution (Wechsler, 1997a), Inspection Time (Deary et al., 2004), and Four-Choice Reaction Time (Deary et al., 2001).
3. **Verbal Memory** – Verbal Paired Associates (Wechsler, 1997b), Logical Memory (Wechsler, 1997b), and Digit Span Backwards (Wechsler, 1997a).
4. **Crystallized Ability** – Verbal Fluency (Lezak et al., 2004), National Adult Reading Test (NART; Nelson & Wilson, 1991), and Wechsler Test of Adult Reading (WTAR; Wechsler, 2001).

All cognitive test scores were standardized (mean = 0, SD = 1) prior to analysis to ensure comparable scaling of factor loadings across measures. General cognitive function (*g*) was estimated using structural equation modelling (SEM) implemented in the lavaan R package (version 0.6.19) (Rosseel, 2012) and the R code for deriving *g* has been previously described (Moodie, Smith, and Cox, 2024). A single latent factor model was specified in which all 13 cognitive tests loaded onto a common latent factor representing *g*. Choice Reaction Time was specified with a negative factor loading so that higher latent *g* scores consistently reflected better cognitive performance across all measures. To account for shared variance among tests within the same cognitive domain that was not attributable to *g*, residual covariances were specified between tests within each cognitive domain. The SEM was fitted using full information maximum likelihood (FIML) estimation to handle missing data under the assumption of missing at random. Individual-level *g* scores were derived as empirical Bayes estimates of the latent factor using the fitted SEM.

### 2.8 Brain MRI measures

MRI data acquisition and processing for the LBC1936 have been detailed previously (Wardlaw et al., 2011). Briefly, participants underwent whole-brain MRI on the same 1.5 T GE Signa Horizon HDx clinical scanner (General Electric, Milwaukee, WI, USA), equipped with a self-shielded gradient system (maximum gradient strength 33 mT/m) and a manufacturer supplied eight-channel phased-array head coil. The structural imaging protocol included whole-brain T1-weighted, T2-weighted, T2*-weighted, and fluid-attenuated inversion recovery (FLAIR) sequences. White matter hyperintensities (WMH) were defined as diffuse, patchy areas in the deep or periventricular white matter, basal ganglia or brain stem, measuring >3 mm in diameter. These regions are hyperintense with respect to normal-appearing grey and white matter on T2-weighted and FLAIR, but not as hyperintense as cerebrospinal fluid (CSF) on T2-weighted images. Quantification of WMH at wave 2 was performed using a semi-automated threshold-based segmentation pipeline that fuses FLAIR and T2*-weighted MRI sequences—after mapping both to the red-green colour space—and applies minimum variance quantisation to the multispectral colour image space to improve tissue discrimination.

Intracranial volume (ICV) was extracted and measured on the T2*-weighted image using the Object Extraction Tool in Analyze 9.0™, as previously described (Zhan et al., 2015). Following ICV extraction, brain tissue segmentation into grey matter, white matter, and CSF was performed on T1-weighted images using FAST (FMRIB’s Automated Segmentation Tool) from the FMRIB Software Library (Zhang, Brady, & Smith, 2001). To minimise misclassification of WMH, FAST-derived tissue segmentations were corrected using WMH binary masks, ensuring that WMH were treated as a distinct tissue class. Further refinement of misclassified dura as normal-appearing white matter was caried out aided by the colour combination of the T1-weighted and T2-weighted images. Regions corresponding to old infarcts were identified using a semi-automated procedure (Hernandez et al., 2012) and excluded from all tissue masks prior to volumetric analyses.

CSF volume comprised all fluid within the cranial cavity, including the ventricles and superficial subarachnoid space. Dura and venous sinuses were separated from the non-brain tissue within the ICV by the T1-weighted-T2-weighted combination mentioned above. Total brain volume (TBV) was defined as ICV minus CSF volume, dura, and venous sinuses. Grey matter volume (GMV) included cortical and deep grey matter and was derived by subtracting white matter, stroke lesions, and WMH masks from the TBV mask. The proportions of TBV, GMV, and WMH volume relative to ICV were calculated and used in statistical analyses to normalise volumetric measures and account for inter-individual differences in head size (Aribisala et al., 2013). All segmentation results were checked for accuracy and manually corrected when/if required.

MRI-derived brain age was predicted using brainageR v2.1 (R code: https://github.com/james-cole/brainageR) (Cole et al., 2018). In brief, T1-weighted MRI scans were segmented into grey matter (GM) and white matter (WM) and subsequently normalised in common space using non-linear spatial registration. The normalised GM and WM images were concatenated and converted into a similarity matrix of training subjects’ data, which was used to predict chronological age using Gaussian Process regression. Model performance was assessed using 10-fold cross-validation, comparing MRI-derived brain age with chronological age (*r* = 0.94). Model coefficients learned during training on the brains of 2,001 healthy adults were then applied to T1-weighted MRI scans acquired from 662 LBC1936 wave 2 participants to predict brain age. The MRI-derived brain age gap was defined as the standardised residual from a linear regression of MRI-derived brain age on chronological age.

### 2.9 Statistical analyses

All analyses were performed using R version 4.4.2 (R Core Team, 2024). ICV–normalised WMH volume, smoking pack years, and alcohol units per week were natural log–transformed [log(x + 1)] due to right skewness and zero inflation. All continuous variables were subsequently standardised (mean = 0, SD = 1). All p-values were adjusted for multiple comparisons using the Benjamini–Hochberg false discovery rate (FDR) procedure as implemented in the stats package in R (Benjamini & Hochberg, 1995; R Core Team, 2024), and results were considered statistically significant at P_FDR_ < 0.05.

#### Correlations

Pairwise correlations among ageing biomarkers were computed using Pearson correlation coefficients with *Hmisc* (version 5.2.2) (Harrell, 2025) and visualised with *corrplot* (version 0.95) (Wei and Simko, 2024).

#### Survival analyses

Cox proportional hazards models examined associations of ageing biomarkers and plasma protein levels, all measured at wave 2, with all-cause mortality in LBC1936 participants followed for up to 17 years, using the survival R package (version 3.8.3) (Therneau, 2024; Therneau and Grambsch, 2000). Time-to-event was defined as the interval between biomarker assessment at wave 2 and the date of death or last follow-up (censoring). Hazard ratios (HRs) reflect the change in mortality risk per one standard deviation increase in the predictor. Biomarkers negatively associated with mortality were labelled with a negative sign in figures, and their reciprocal HRs (1/HR) were plotted for effect size comparability. Proportional hazards assumptions were evaluated through Schoenfeld residuals using the cox.zph function from the survival package. For each association failing to meet the assumption (Schoenfeld residuals p < 0.05), a sensitivity analysis was run across yearly follow-up intervals (Supplementary Figures 3 & 4).

We fitted separate Cox models for each ageing biomarker, adjusted for chronological age and sex, as well as a model including all biomarkers adjusted for chronological age, sex, and modifiable risk factors (smoking, alcohol consumption, and BMI). Highly collinear biomarker pairs (Pearson’s r > 0.7, P_FDR_ < 0.05; FER–FEV and GMV–TBV) were handled by randomly excluding one biomarker from each pair, resulting in 21 biomarkers in the saturated model.

Plasma protein associations with mortality were assessed using separate Cox models for each protein, with incremental adjustment for chronological age, sex, kidney function (estimated glomerular filtration rate, eGFR), and modifiable risk factors. eGFR was calculated from serum creatine levels, chronological age, and sex according to the CKD-EPI Creatinine Equation (National Kidney Foundation, 2021; Inker et al., 2021). To address the high dimensionally and multicollinearity in proteomics, we further ran an elastic net–penalised Cox regression on all 8,699 proteins simultaneously using the *glmnet* R package (version 4.1.8) (Friedman, Tibshirani, and Hastie, 2010; Tay, Narasimhan, and Hastie, 2023). The mixing parameter (α) was set to 0.5, and the penalty parameter (λ) was chosen as 0.16 based on the λ_min_ from 5-fold cross-validation. Alive and dead subjects were evenly distributed across folds. Detailed model specifications are provided in Table 2.

**TABLE 2.** Description of survival analysis models.

| Model | Description | Predictors | Covariates | Figure |
| --- | --- | --- | --- | --- |
| Single ageing biomarker model | Separate Cox regression for each ageing biomarker | Single biomarker | Chronological age and sex | 1A |
| Multiple ageing biomarker model | Single Cox regression including all selected biomarkers | 21 biomarkers | Chronological age, sex, BMI, smoking pack years, and alcohol units per week | 1B |
| Basic proteomic model | Separate Cox regression for each protein | Single protein | Chronological age and sex | 2A |
| Kidney-adjusted proteomic model | Separate Cox regression for each protein | Single protein | Chronological age, sex, and eGFR | 2A |
| Fully adjusted proteomic model | Separate Cox regression for each protein | Single protein | Chronological age, sex, eGFR, BMI, smoking pack years, and alcohol units per week | 2A, 2B |
| Penalised proteomic model | Elastic net-penalised Cox regression including all proteins for mortality risk prediction | 8,699 proteins | None | 3 |

#### Enrichment analyses

Proteins that were significantly associated with mortality in the fully adjusted Cox models were subjected to enrichment analyses. Gene Ontology Biological Process enrichment was performed using *gProfiler2* (version 0.2.4) (Kolberg et al., 2020), with all 8,699 proteins used as the background. Tissue-specific expression enrichment was conducted using the GTEx (Genotype-Tissue Expression) database, implemented through the *TissueEnrich* R package (version 1.26.0) (Jain and Tuteja, 2018).

## 3 Results

### 3.1 Characterising organ ageing signatures in the LBC1936

We predicted ages for eleven major organs in LBC1936 wave 2 participants (Supplementary Figure 1A); organ age gaps were calculated as the residuals from linear regressions of predicted organ age on chronological age and were standardised to have zero mean and unit variance (Supplementary Figure 1B). Although LBC1936 wave 2 participants were of similar chronological age (range: 71.0–74.2 years), considerable variability in organ ageing (55.2–99.6 years) was observed (Supplementary Figure 1A). Pairwise Pearson correlation analysis revealed weak to moderate associations between organ age gaps (r = 0.08–0.57, P_FDR_ < 0.05) (Supplementary Figure 1C). The strongest correlations were observed between adipose tissue and the immune system (r = 0.57), adipose tissue and the pancreas (r = 0.55), and the immune system and the liver (r = 0.55), which may reflect partially shared ageing patterns and are consistent with their functional overlap in metabolic and immune processes. Next, we quantified extreme organ ageing, defined as an organ age gap greater than 1.5 SD above the mean. Most participants (62.5%, N=497) exhibited no extreme organ ageing; 19.9% (N=158) showed extreme ageing in one organ, 7.7% (N=61) in two organs, and 3.8% (N=30) in five or more organs (multi-organ agers), indicating that concurrent extreme ageing across multiple organs is uncommon in this population (Supplementary Figure 1D).

### 3.2 Epigenetic, neuroimaging, and functional ageing biomarkers outperform proteomic organ ages in associations with all-cause mortality

We benchmarked these eleven proteomic organ age measures against other multimodal ageing biomarkers (measured at wave 2) for associations with all-cause mortality in the LBC1936. Separate Cox proportional hazards models were fitted for each biomarker, adjusted for chronological age and sex (Figure 1A). All ageing biomarkers were significantly associated with mortality (HR_range_ = 1.11–1.62, P_FDR_ < 0.05), except telomere length. Among the proteomic organ age measures, a 1-SD increase in each organ age corresponded to a 16–43% higher mortality hazard. The most pronounced effects were observed for accelerated ageing of the liver (HR = 1.43, 95% CI = 1.30–1.58, P_FDR_ = 8.00×10^-13^), immune system (HR = 1.42, 95% CI = 1.29–1.57, P_FDR_ = 1.79×10^-11^), and heart (HR = 1.38, 95% CI = 1.25–1.53, P_FDR_ = 2.58×10^-10^). The proteomic brain age gap (HR = 1.31, 95% CI = 1.19–1.44, P_FDR_ = 1.27×10^-7^) showed a similar hazard ratio to the MRI-derived brain age gap (HR = 1.30, 95% CI = 1.15–1.46, P_FDR_ = 1.88×10^-5^). However, only the MRI-derived brain age gap was associated with neuroimaging measures, with greater brain age acceleration correlating with lower TBV (r = −0.47) and GMV (r = −0.43), and higher WMH volume (r = 0.19) (Supplementary Figure 2).

**FIGURE 1.**
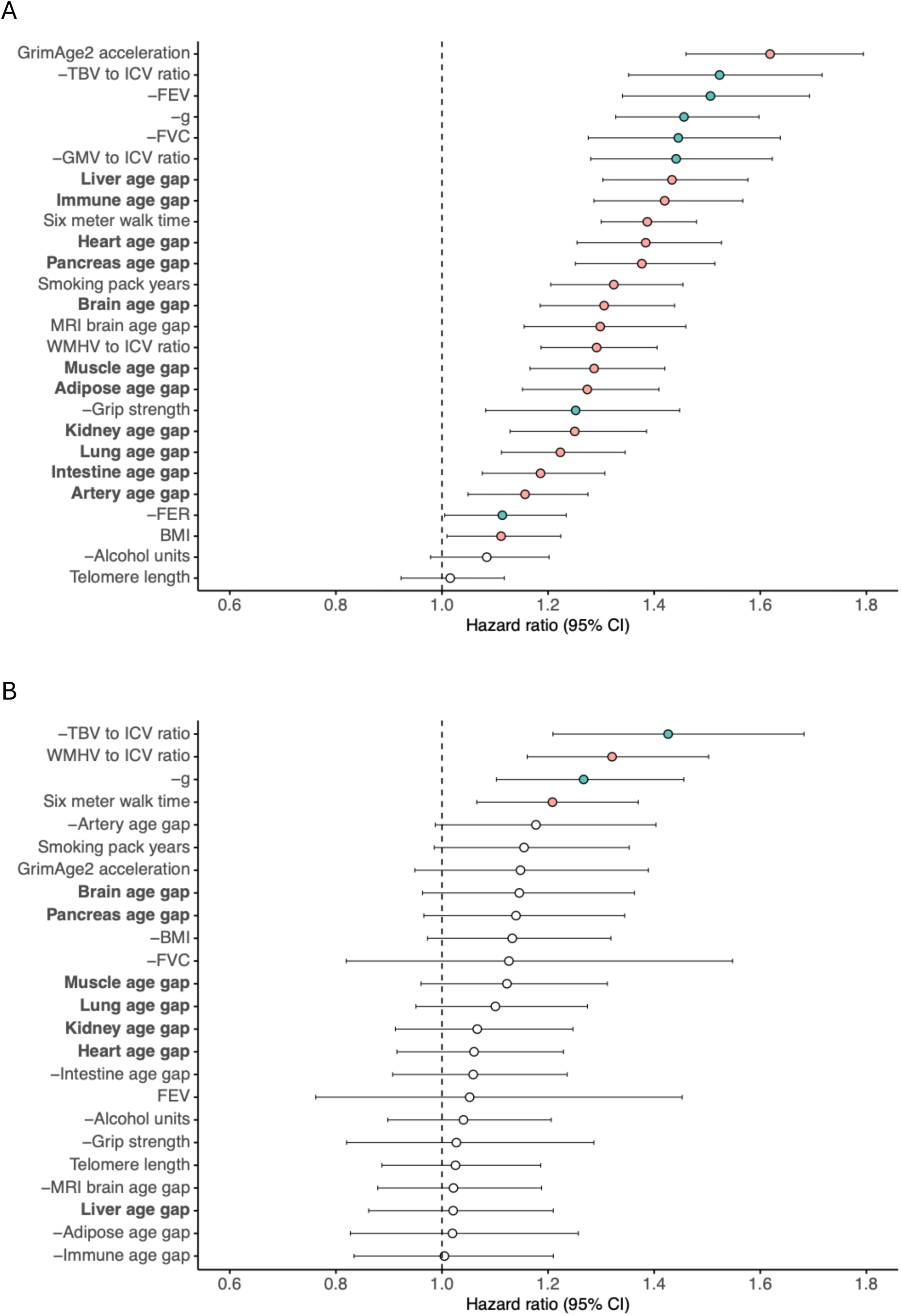
Associations of ageing biomarkers with all-cause mortality in the LBC1936. **(A)** Associations of ageing biomarkers with all-cause mortality were estimated by fitting separate Cox proportional hazards models for each biomarker, adjusted for chronological age and sex (n = 326–444 deaths in 656–861 individuals, varying by biomarker). **(B)** Results from a single Cox proportional hazards model including all ageing biomarkers as predictors of all-cause mortality, adjusted for chronological age, sex, and modifiable risk factors, including smoking pack-years, alcohol consumption, and body mass index (BMI) (n = 237 deaths in 460 individuals). Grey matter volume (GMV) and forced expiratory ratio (FER) were excluded due to high collinearity with total brain volume (TBV) and forced expiratory volume (FEV), respectively. Biomarkers negatively associated with mortality are shown in green, and biomarkers positively associated with mortality are highlighted in pink. All negatively associated biomarkers are labelled with a minus sign preceding their name, and their reciprocal hazard ratios (1/HR) are plotted to facilitate comparison of effect sizes. The vertical dotted line indicates a hazard ratio of 1. Circles represent estimated hazard ratios, with error bars showing 95% confidence intervals. P-values were adjusted for multiple comparisons using the Benjamini–Hochberg (BH) false discovery rate (FDR). Non-significant p-values are shown in white, while significant values (P_FDR_ < 0.05) are highlighted in colour. Proteomic organ age gaps are indicated in bold.

Several other ageing biomarkers were more strongly associated with mortality than proteomic organ ages (Figure 1A). GrimAge2 acceleration demonstrated the largest overall association with mortality, with a 1-SD increase conferring a 62% higher mortality hazard (HR = 1.62, 95% CI = 1.46–1.79, P_FDR_ = 7.67×10^-19^). Moreover, smaller TBV (reciprocal HR = 1.52, 95% CI = 1.35–1.72, P_FDR_ = 2.14×10^-11^) and GMV (reciprocal HR = 1.44, 95% CI = 1.28–1.62, P_FDR_ = 3.75×10^-9^), reduced respiratory function (FEV: reciprocal HR = 1.51, 95% CI = 1.34–1.69, P_FDR_ = 2.39×10^-11^; FVC: reciprocal HR = 1.45, 95% CI = 1.28–1.64, P_FDR_ = 1.47×10^-8^), and poorer cognitive performance (reciprocal HR = 1.46, 95% CI = 1.33–1.60, P_FDR_ = 1.58×10^-14^) were each associated with a 44–52% higher mortality risk per 1-SD decrease in their levels. Domain-specific trends revealed that GrimAge2 acceleration was the strongest predictor among blood-based biomarkers, TBV among neuroimaging markers, FEV among physical function measures, and smoking among modifiable risk factors. Detailed results, including hazard ratios, confidence intervals, p-values, and proportional hazards (PH) assumption violations, are provided in Supplementary Table 1 and summarised in Figure 1A. For ageing biomarkers that violated the PH assumption, plots of hazard ratios per year of follow-up are included in Supplementary Figure 3 to demonstrate that hazard ratios remain largely stable, including instances where the assumptions are met, apart from the first few years when the incidence of death was low.

### 3.3 Neuroimaging, cognitive, and physical function biomarkers of ageing independently associate with all-cause mortality

To evaluate the independent contributions of ageing biomarkers to all-cause mortality, we modelled all biomarkers in a single multivariable Cox proportional hazards regression model adjusted for chronological age, sex, and modifiable risk factors (smoking, alcohol consumption, and BMI) (Figure 1B). FER and GMV, which showed high collinearity (Pearson’s r > 0.7, P_FDR_ < 0.05) with FEV and TBV, respectively, were excluded from this analysis (Supplementary Figure 2). Only four biomarkers—TBV, WMH volume, *g*, and walk time—retained significant independent associations with mortality. Notably, proteomic organ ageing biomarkers did not show independent associations with mortality, suggesting that their relationships may be largely mediated by or confounded with these four core measures. Collectively, TBV, WMH volume, *g*, and walk time captured a substantial proportion of mortality risk beyond basic demographic factors (chronological age and sex), increasing the explained variance roughly seven-fold, from 3% to 20%, as measured by Nagelkerke pseudo-R². Hazard ratios, confidence intervals, p-values, and PH assumption violations are listed in Supplementary Table 2, and the corresponding forest plot is depicted in Figure 1B.

### 3.4 Plasma proteomic signatures, biological processes, and tissues associated with all-cause mortality

To identify proteomic biomarker candidates of mortality risk and to characterise their underlying biological functions and tissue origins, we conducted a large-scale survival analysis of SomaScan 11K plasma proteins (8,699 unique UniProt IDs) in the LBC1936. Chronological age- and sex-adjusted Cox proportional hazards regression identified 368 proteins significantly associated with mortality (P_FDR_ < 0.05) (Figure 2A). After additional adjustment for kidney function (eGFR), 294 protein–mortality associations remained. These were further reduced to 202 associations after adjusting for modifiable risk factors (smoking, alcohol consumption, and BMI). In this fully adjusted Cox model, the strongest positive associations with mortality were observed for GDF15 (growth differentiation factor 15, HR = 1.53, 95% CI = 1.37–1.72, P_FDR_ = 1.04×10^-9^), CST3 (cystatin C, HR = 1.48, 95% CI = 1.29–1.69, P_FDR_ = 1.34×10^-5^), and COL18A1 (Collagen Type XVIII Alpha 1 Chain, HR = 1.47, 95% CI = 1.30–1.68, P_FDR_ = 6.15×10^-6^) (Figure 2B). Conversely, HPGDS (hematopoietic prostaglandin D synthase, reciprocal HR = 1.38, 95% CI = 1.22–1.56, P_FDR_ = 9.83×10^-5^), NPS (neuropeptide S, reciprocal HR = 1.35, 95% CI = 1.22–1.49, P_FDR_ = 1.27×10^-5^), and BAGE3 (B melanoma antigen 3, reciprocal HR = 1.34, 95% CI = 1.21–1.49, P_FDR_ = 1.75×10^-5^) showed the strongest negative associations with mortality. The top five mortality-associated proteins (HR_range_ = 1.45–1.53) had higher hazard ratios than all the proteomic organ age gaps (HR_range_ = 1.16–1.43), but lower than GrimAge2 acceleration (HR = 1.62) (Figure 1A). Only 23 of the 202 mortality-associated proteins were included in the organ age models, indicating minimal overlap between the two sets. Gene ontology biological process enrichment of the 202 mortality-associated proteins revealed enrichment in multiple immune processes (namely, antimicrobial humoral response, chemotaxis, and cell adhesion) as well as extracellular matrix organisation (Figure 2C). Furthermore, tissue enrichment analysis identified the liver as the only significantly enriched tissue based on GTEx bulk tissue RNA sequencing data (Figure 2D). Hazard ratios, confidence intervals, p-values, and PH assumption violations for proteins significantly associated to mortality are reported in Supplementary Table 3 and annual plots of hazard ratios and PH assumption violations for the top 20 proteins are shown in Supplementary Figure 4.

**FIGURE 2.**
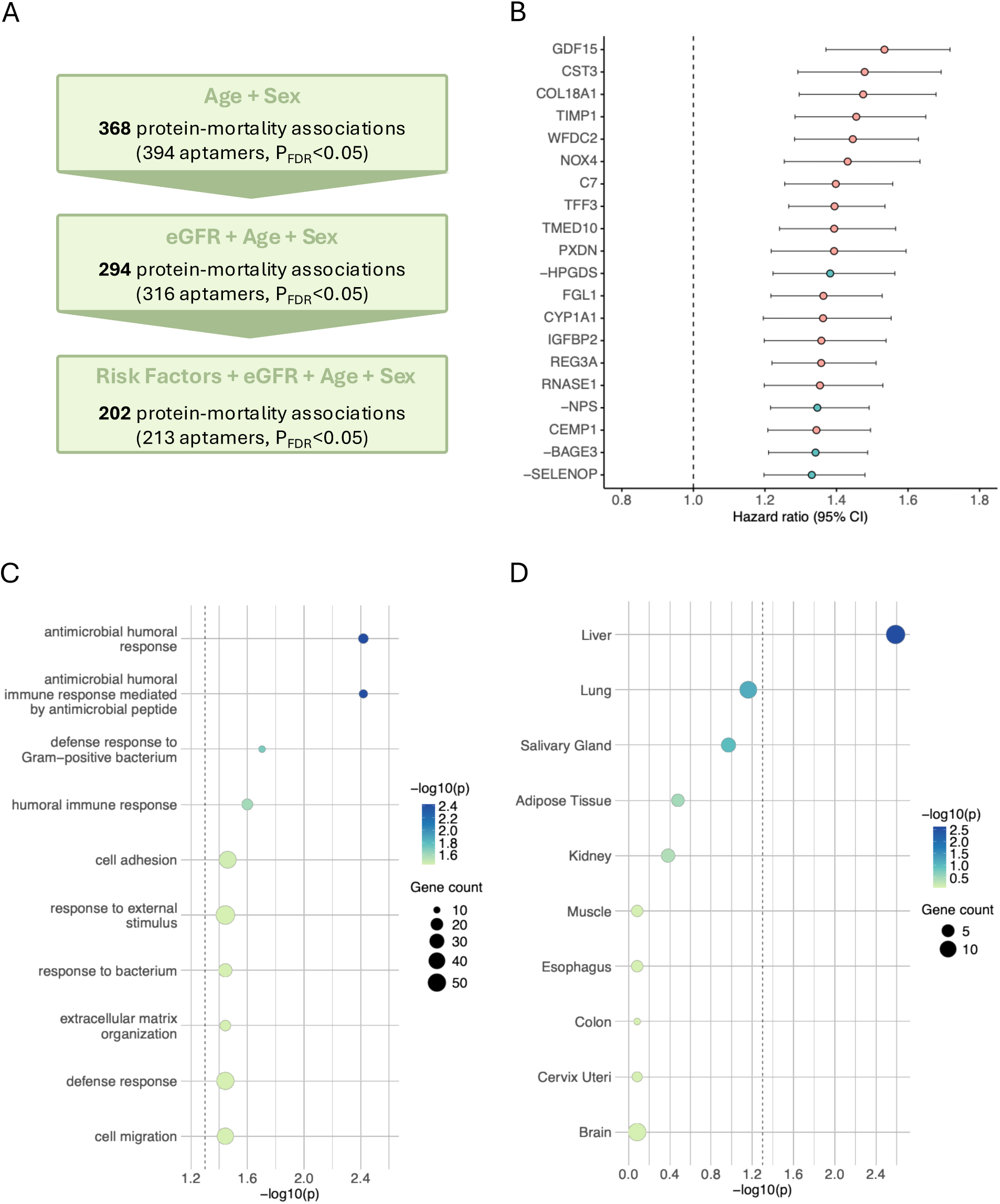
Plasma proteomic associations with all-cause mortality and enriched biological processes and tissues in the LBC1936. **(A)** Associations of SomaScan 11K plasma proteins with all-cause mortality. Cox proportional hazards models were fitted separately for each protein. Basic models adjusted for chronological age and sex identified 368 significant protein–mortality associations. Additional adjustment for kidney function, measured by estimated glomerular filtration rate (eGFR), yielded 294 significant associations, and fully adjusted models including modifiable risk factors (smoking, alcohol consumption, and body mass index [BMI]) retained 202 significant associations. P-values were adjusted for multiple comparisons using the Benjamini–Hochberg (BH) false discovery rate (FDR). **(B)** Top 20 significant protein–mortality associations from the fully adjusted Cox model, ranked by the largest hazard ratios (n = 358 deaths in 694 individuals). Proteins negatively associated with mortality are shown in green, and proteins positively associated with mortality are highlighted in pink. All negatively associated proteins are labelled with a minus sign preceding their name, and their reciprocal hazard ratios (1/HR) are plotted to facilitate comparison of effect sizes. Circles represent estimated hazard ratios, with error bars showing 95% confidence intervals. The dotted line indicates a hazard ratio of 1. **(C)** Gene Ontology biological process enrichment of the 202 significant mortality-associated proteins identified in the fully adjusted Cox model. **(D)** Tissue enrichment of the same 202 proteins based on Genotype-Tissue Expression (GTEx) bulk tissue RNA-sequencing data. The dotted line indicates significance at −log_10_(P_FDR_) > 1.3 (equivalent to P_FDR_ < 0.05).

To account for the high dimensionality and multicollinearity of plasma proteomic data, we fitted an elastic net–penalised Cox proportional hazards regression model simultaneously to 8,699 SomaScan 11K plasma proteins measured in LBC1936 wave 2 participants (5-fold cross-validation; α = 0.5; λ_min_ = 0.16). In this model, each protein’s coefficient represents the log hazard ratio for a 1-SD increase in its plasma level, with positive values indicating higher mortality risk and negative values indicating lower risk. The model retained 37 proteins with non-zero coefficients; GDF15 (coefficient = 0.18, HR ≈ 1.20) had the largest positive coefficient, while HPGDS (coefficient = −0.09, HR ≈ 0.91) had the largest negative (protective) coefficient (Figure 3), consistent with results from the fully adjusted individual protein-mortality Cox models (Figure 2B). Moreover, 32 of the 37 elastic net proteins (86.5%) overlapped with those identified by the fully adjusted individual protein-mortality Cox models, indicating strong concordance between the two approaches. Estimated elastic net model protein coefficients are provided in Supplementary Table 4.

**FIGURE 3.**
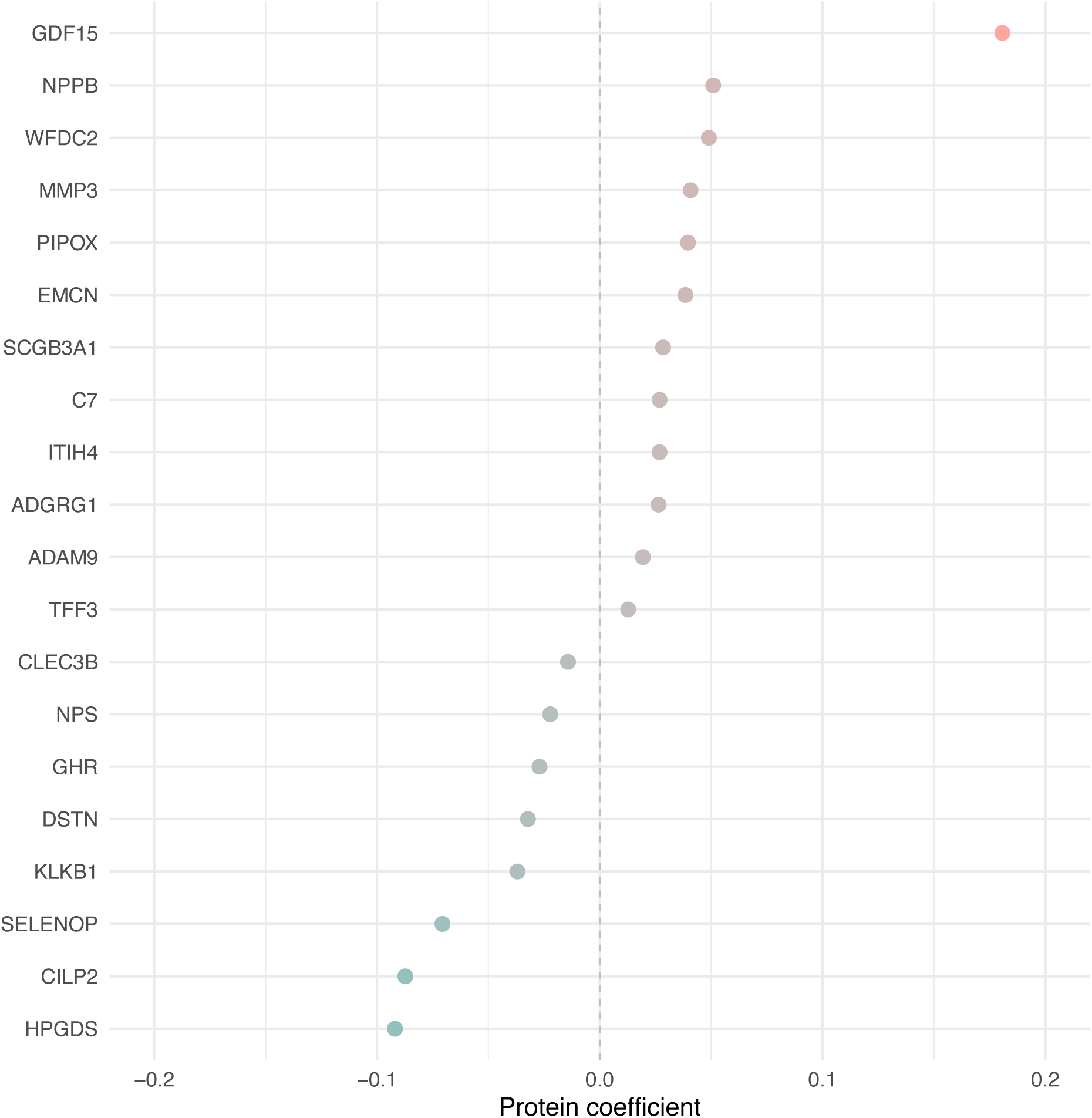
Top 20 plasma protein predictors of all-cause mortality in the LBC1936. The plot shows the 20 SomaScan 11k plasma proteins with the largest coefficients from a total of 37 proteins selected to predict all-cause mortality using elastic net–penalised Cox regression with 5-fold cross-validation (n = 404 deaths in 789 individuals). The mixing parameter (α) was set to 0.5, and the penalty parameter (λ) was chosen as 0.16 based on the λ_min_ from 5-fold cross-validation. Proteins with positive coefficients, indicating higher mortality risk, are shown in pink, while proteins with negative coefficients, indicating lower mortality risk, are shown in green.

## 4 Discussion

This study benchmarked eleven plasma proteomic organ clocks against other multimodal ageing biomarkers in relation to all-cause mortality over up to 17 years of follow-up in the LBC1936. Accelerated organ ageing conferred a 16–43% higher mortality risk per 1-SD increase, with the liver, immune system, and heart showing the largest effects. However, greater GrimAge2 acceleration, smaller MRI-derived TBV and GMV, reduced respiratory function (FEV and FVC), and lower *g* exhibited larger hazard ratios than any organ age, increasing mortality risk by 44–62%. In the model including all biomarkers simultaneously, only TBV, WMH volume, *g*, and walk time remained significantly associated with mortality. Survival analyses of 8,699 SomaScan 11K plasma proteins identified 202 proteins significantly associated with mortality. These proteins were primarily enriched for immune-related biological processes and were predominantly expressed in the liver. GDF15, CST3, and COL18A1 showed the strongest positive associations with mortality risk, increasing it by 47–53% per 1-SD increase in plasma levels. In contrast, HPGDS, NPS, and BAGE3 were the top negatively associated proteins, increasing mortality risk by 34–38% per 1-SD decrease in plasma levels. A penalised regression model with all proteins as inputs selected 37 candidates in a parsimonious solution, with GDF15, NPPB, and WFDC2 among the most influential predictors. Overall, this study comprehensively benchmarks proteomic organ ages against other multimodal ageing biomarkers and uncovers new plasma proteomic signatures and biological pathways associated with mortality.

### 4.1 Comparative utility of proteomic organ ages relative to other ageing biomarkers for mortality risk assessment

Our study confirms that accelerated proteomic organ ageing is associated with increased mortality risk in an independent longitudinal cohort of older adults. Our findings align with the original study, which examined associations between the eleven organ ages and 15-year all-cause mortality risk in the LonGenity cohort (N=864) using Cox proportional hazards regression (Oh et al., 2023). The LonGenity study reported a 17–53% increase in mortality risk per 1-SD increase in organ age, compared with a 16–43% increase observed in our study. Here, however, we demonstrated that organ ages perform modestly relative to other ageing biomarkers, including GrimAge2 acceleration, brain MRI, respiratory function, and cognitive measures. GrimAge2 has previously been identified as the strongest predictor of 10-year all-cause mortality among 14 widely used epigenetic clocks in the Generation Scotland cohort (N=18,859) (Mavrommatis et al., 2025). Its superior performance in the present study may be due to its ability to capture systemic ageing biology and from it being trained on mortality outcomes.

The finding that TBV, WMH volume, cognitive performance, and walk time remained independently associated with mortality—even after adjustment for a wide range of ageing biomarkers—suggests that these measures are among the most informative candidates for multimodal mortality risk stratification. Notably, proteomic organ ageing biomarkers did not retain independent associations with mortality, suggesting that their relationships may be largely mediated by or confounded with these four core measures. Our effect sizes are consistent with prior work showing that MRI-derived brain structural measures (Debette et al., 2019; Ghaznawi et al., 2021; Van Elderen et al., 2016) and walking speed (Abellan van Kan et al., 2009; Peel et al., 2018; Studenski et al., 2011) are robust predictors of survival. Lower ICV–normalised TBV was associated with a 52% higher all-cause mortality risk in the AGES-Reykjavik study (Van Elderen et al., 2016), an effect size identical to that observed in LBC1936. Similarly, greater ICV–adjusted WMH volume conferred a 32% higher mortality risk in SMART-MR study (Ghaznawi et al., 2021), closely matching the 29% increase observed in LBC1936.

### 4.2 Mortality-associated proteins have key immune-related functions

Mortality-associated proteins identified in our study are enriched in the liver and in immune-related biological processes, underscoring the central role of immune function in longevity. Notably, this aligns with our findings that accelerated liver and immune ageing show the strongest associations with mortality among the proteomic organ age measures. Ageing is accompanied by a progressive decline in immune function (immunosenescence), which includes impaired phagocytosis, chemotaxis, and natural killing by innate immune cells; reduced lymphocyte production and activation; and the emergence of a senescence-associated secretory phenotype (SASP) that drives low-grade chronic inflammation (Cisneros et al., 2022; Liu et al., 2023; Lynch et al., 2009; Theodorakis et al., 2024). These deficits increase susceptibility to infection and weaken immune defence, contributing to higher mortality from infectious diseases in older age (Du et al., 2021; Ittisanyakorn et al., 2019; Theodorakis et al., 2024). Consistent with this, several of the most overrepresented mortality-associated biological processes in our study are related to the humoral immune response, the branch of adaptive immunity in which B lymphocytes produce antibodies that neutralize extracellular pathogens, including bacteria and viruses (Janewa et al., 2001).

GDF15 (growth differentiation factor 15), a key stress-, infection-, and inflammation-induced cytokine regulating immune responses (Pence, 2022), emerged as the strongest individual protein predictor of mortality in the LBC1936. Circulating GDF15 levels increase with age, acute or chronic illness, and several age-related diseases—including cardiovascular disease, type 2 diabetes, neurodegeneration, and cancer—and consistently predict higher all-cause mortality across cohorts (Bao et al., 2021; Conte et al., 2022; Gadd et al., 2024; Lajer et al., 2010; Lyu et al., 2024). TFF3 (Trefoil factor 3), which maintains mucosal integrity and modulates immune responses at epithelial barriers, was also positively associated with mortality in our study. Notably, TFF3 is co-upregulated with GDF15 in heart failure and likewise predicts increased mortality risk in peripheral artery disease patients (Ceelen et al., 2022; Pesau et al., 2024). Another mortality-associated protein was CST3 (cystatin C) a cysteine protease inhibitor involved in antigen processing, cytokine secretion, nitric oxide synthesis, apoptosis, and host defence (Turk et al., 2002; Zi et al., 2018). Dysregulated CST3 expression is implicated in inflammatory autoimmune pathologies and tumour development, while elevated serum levels associate with higher all-cause mortality in coronary heart disease patients (Ix et al., 2007). Complement component C7, a key constituent of the membrane attack complex, has similarly been associated with increased mortality risk in patients with suspected acute coronary syndrome (Aarsetøy et al., 2021). Finally, REG3A (Regenerating islet-derived protein 3 alpha), a gut-expressed C-type lectin involved in antibacterial innate host defence and immunomodulation, also emerged as a mortality marker, consistent with prior observations linking elevated plasma REG3A to adverse outcomes (Faivre et al., 2025; Sands et al., 2024).

Immune-related proteins negatively associated with mortality risk in the LBC1936 included HPGDS (hematopoietic prostaglandin D synthase), which synthesises the bioactive lipid mediator prostaglandin D_2_ that regulates inflammatory responses and allergic reactions (Arima et al., 2011; Trivedi et al., 2006), and NPS (neuropeptide S), which reduces anxiety, enhances memory, and regulates metabolism and immune functions, such as macrophage phagocytosis and monocyte chemotaxis (Filaferro et al., 2013; Pape et al., 2010; Pulkkinen et al., 2006; Reinscheid, 2008; Zhao et al., 2019). SELENOP (selenoprotein P), a liver-derived selenium transporter that protects against oxidative stress and regulates inflammation, also emerged as protective in the LBC1936. Low SELENOP plasma levels have also been associated with higher all-cause and cardiovascular mortality in other cohorts (Hariharan et al., 2020; Huang et al., 2012; Schomburg et al., 2019). Collectively, these findings emphasise the critical role of immune-related proteins in shaping mortality risk in older adults.

### 4.3 Mortality-associated proteins are involved in extracellular matrix organisation

Another overrepresented biological function among mortality-associated proteins is extracellular matrix (ECM) organisation. Many mortality-associated proteins are either structural components of the ECM or regulators of ECM dynamics, including several collagen proteins (COL4A5, COL6A3, and COL18A1) that contribute to ECM structure within basement membranes or the pericellular matrix (Cosgrove and Liu, 2017; Heljasvaara et al., 2017; Marneros and Olsen, 2005; Wang and Pan, 2020); TIMP1, an inhibitor of matrix metalloproteinases (MMPs) that modulates ECM turnover, composition, and tissue remodelling (Brew et al., 2010); PXDN, a peroxidase that contributes to ECM assembly and stabilisation by catalysing collagen IV crosslinking in basement membranes (Lázár et al., 2015); ANTXR2, which is essential for ECM remodelling via MT1-MMP activation and for regulation of collagen VI (Metti et al., 2025; Reeves et al., 2012); and WFDC2, an activator of MMP-2, which is a key enzyme in ECM degradation (Chen et al., 2022). The ECM provides structural integrity, tissue elasticity, controlled diffusion of nutrients and metabolites, and biomechanical cues essential for cellular adhesion, migration, proliferation, differentiation, and overall tissue homeostasis (Birch et al., 2019; Bosman et al., 2003). Age-related biochemical modifications of long-lived ECM proteins—including glycation, oxidation, fragmentation, and aberrant cross-linking—reduce tissue elasticity, increase matrix stiffness, impair ECM-mediated signalling, and promote inflammatory responses that drive fibrosis (Birch et al., 2018; Gautieri et al., 2016; Vijg et al., 2008; Wick et al., 2013). ECM deterioration has been mechanistically implicated in multiple age-related diseases, including cardiovascular disease, diabetes, atherosclerosis, neuropathies, and cancer, underscoring its central role in multisystem pathology (Boaru et al., 2025; Del Turco et al., 2012; Snedeker and Gautieri, 2014; Statzer et al., 2023; Vincent et al., 2004; Vijg et al., 2008). Consequently, fibrotic complications are estimated to contribute to approximately 45% of deaths (Henderson et al., 2020). In addition, circulating fragments of degraded ECM, such as elastin-derived peptides, can actively promote ageing through activation of innate immune pathways, reducing healthspan and lifespan in animal models (Yi et al., 2025). Overall, age-dependent ECM deterioration is an emerging, yet understudied, hallmark of ageing and longevity that warrants further research.

### 4.4 Limitations

Although this study has notable strengths, including a broad range of ageing biomarkers, large-scale unbiased proteomic profiling, and direct effect-size comparisons within a single population, several limitations should be considered. First, LBC1936 consists of relatively healthy, ethnically homogeneous Scottish individuals of similar chronological age, which may limit the generalisability of our findings to more diverse populations. Second, our analyses relied on baseline biomarker values and did not incorporate longitudinal trajectories, which may provide greater predictive value than single time-point measurements. Future analyses will consider protein trajectories with health and survival. Third, due to sample size, we examined all-cause rather than cause-specific mortality, which may be more informative for organ age biomarkers. Fourth we used a single proteomics platform (SomaScan 11K) and were therefore dependent on the reliability of those protein measurements.

### 4.5 Conclusion

This study benchmarks proteomic organ ages against a broad array of multimodal ageing biomarkers for predicting all-cause mortality risk. Although accelerated ageing in all organs is linked to higher mortality, organ age measures exhibit smaller hazard ratios than epigenetic age acceleration, structural brain MRI, respiratory function, and cognitive biomarkers, and they do not retain independent predictive value when all ageing biomarkers and modifiable risk factors are accounted. The possibility that they offer enhanced risk prediction for specific mortality causes remains unknown. Additionally, our large-scale, unbiased proteomic analysis identified novel protein candidates that predict all-cause mortality and highlighted their roles in immune regulation and extracellular matrix organisation. These proteins were largely independent of those that contributed to the proteomic organ age models. External validation in independent cohorts will be necessary to assess the predictive performance of these biomarkers for mortality risk stratification and their potential utility as surrogate endpoints in longevity trials.

## Author Contributions

M.P., R.E.M., and S.E.H. designed the study. M.P. conducted all data analyses and wrote the original manuscript draft. I.M.E. performed quality control of the SomaScan 11K plasma proteomic data for LBC1936 wave 2. P.R. and J.C. were responsible for data wrangling and quality assurance as well as for data collection, linkage and mortality follow-up in the LBC1936. M.V.H. and J.M.W. contributed to the brain MRI methods section. All authors provided critical feedback, reviewed, and edited the manuscript.

## Acknowledgments

The authors thank all LBC1936 study participants and research team members who have contributed, and continue to contribute, to ongoing studies. They also thank the nursing and Genetics Core staff at the Wellcome Trust Clinical Research Facility, Western General Hospital, Edinburgh, and the staff at the National Records of Scotland for their support.

## Funding

The Lothian Birth Cohorts received joint funding from the BBSRC and ESRC (BB/W008793/1, to S.R.C., which supported S.E.H., J.C., and P.R.), the MRC (MR/R024065/1), Age UK (The Disconnected Mind), the Milton Damerel Trust, and the University of Edinburgh. DNA methylation profiling was funded by the Centre for Cognitive Ageing and Cognitive Epidemiology (Pilot Fund Award), Age UK, the Wellcome Trust Institutional Strategic Support Fund, The University of Edinburgh, and The University of Queensland. M.P. was supported by the Translational Neuroscience PhD Programme, funded by the Wellcome Trust (218493/Z/19/Z). S.R.C was supported by a Sir Henry Dale Fellowship, jointly funded by the Wellcome Trust and the Royal Society (221890/Z/20/Z). S.E.H., S.R.C, and R.E.M. were additionally funded by the National Institutes of Health (NIH) research grant (U01AG083829), which also supported I.M.E., T.C.R., I.J.D., E.M.T.D., M.V.H., and by the BBSRC grant (UKRI1941). J.M.W. was supported by the UK DRI, which is funded by the MRC, Alzheimer’s Society and Alzheimer’s Research UK, as well as the NIH. M.V.H. received funding from the Row Fogo Charitable Trust (BRO-D.FID3668413, to J.M.W.). This work was also supported in part by the National Institute on Aging (NIA) Intramural Research Program (K.A.W.). The contributions of the NIH author(s) are considered Works of the United States Government. The findings and conclusions presented in this paper are those of the author(s) and do not necessarily reflect the views of the NIH or the U.S. Department of Health and Human Services.

## Ethics

LBC1936 ethical approval was obtained from the Scotland A Research Ethics Committee (07/MRE00/58). All participants provided written informed consent.

## Conflicts of Interest

R.E.M is an advisor to the Epigenetic Clock Development Foundation and Optima Partners Ltd. K.A.W. is an Associate Editor for Alzheimer’s & Dementia: The Journal of the Alzheimer’s Association, Alzheimer’s & Dementia: Translational Research and Clinical Interventions (TRCI), and on the Editorial Board of Annals of Clinical and Translational Neurology. K.A.W. is on the Board of Directors of the National Academy of Neuropsychology. K.A.W. has given unpaid presentations and seminars on behalf of SomaLogic. K.A.W. is a co-founder of Centia Bio. The work presented in this manuscript was conducted independently of Centia Bio and without financial support from the company. The remaining authors have no conflicts of interest to disclose.

## Data Availability Statement

All data produced in the present study are reported in Supplementary Tables. The code used for analyses is openly accessible at the following GitHub repository: https://github.com/marioni-group/Mortality_Biomarkers_LBC1936.git. The original data analysed in this study are not publicly available because they contain sensitive information that could compromise participant consent and confidentiality. Data access instructions for the LBC are available at: https://www.ed.ac.uk/lothian-birth-cohorts/data-access-collaboration.

---

**SUPPLEMENTARY FIGURE 1.**
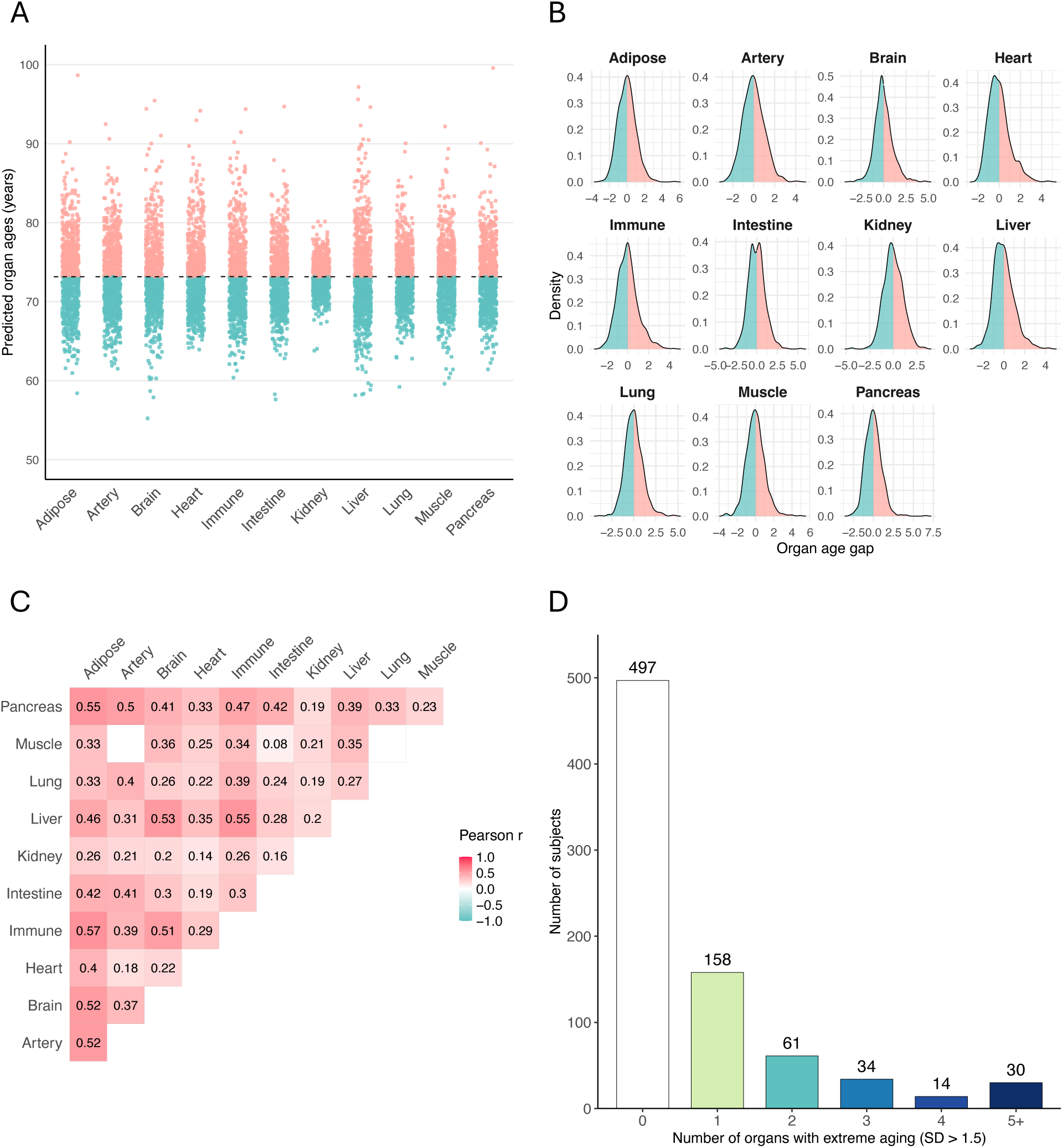
Characterisation of plasma proteomic organ ageing signatures in the LBC1936. **(A)** Predicted ages for eleven organs in LBC1936 wave 2 participants (N = 795). Each point represents an individual, and the black dashed line indicates the mean organ age. **(B)** Density plots of organ age gaps, defined as the standardised residuals (mean = 0, SD = 1) from the linear regression of predicted organ age on chronological age. Positive values, indicating accelerated ageing, are shown in pink, and negative values, indicating slower ageing, are shown in green. **(C)** Pairwise Pearson correlation matrix of organ age gaps; only significant correlations with P_FDR_ < 0.05 are displayed. Positive correlations are shown in pink, and negative correlations in green. **(D)** Number of individuals exhibiting extreme organ ageing (standard deviation > 1.5) in a single or multiple organs.

**SUPPLEMENTARY FIGURE 2.**
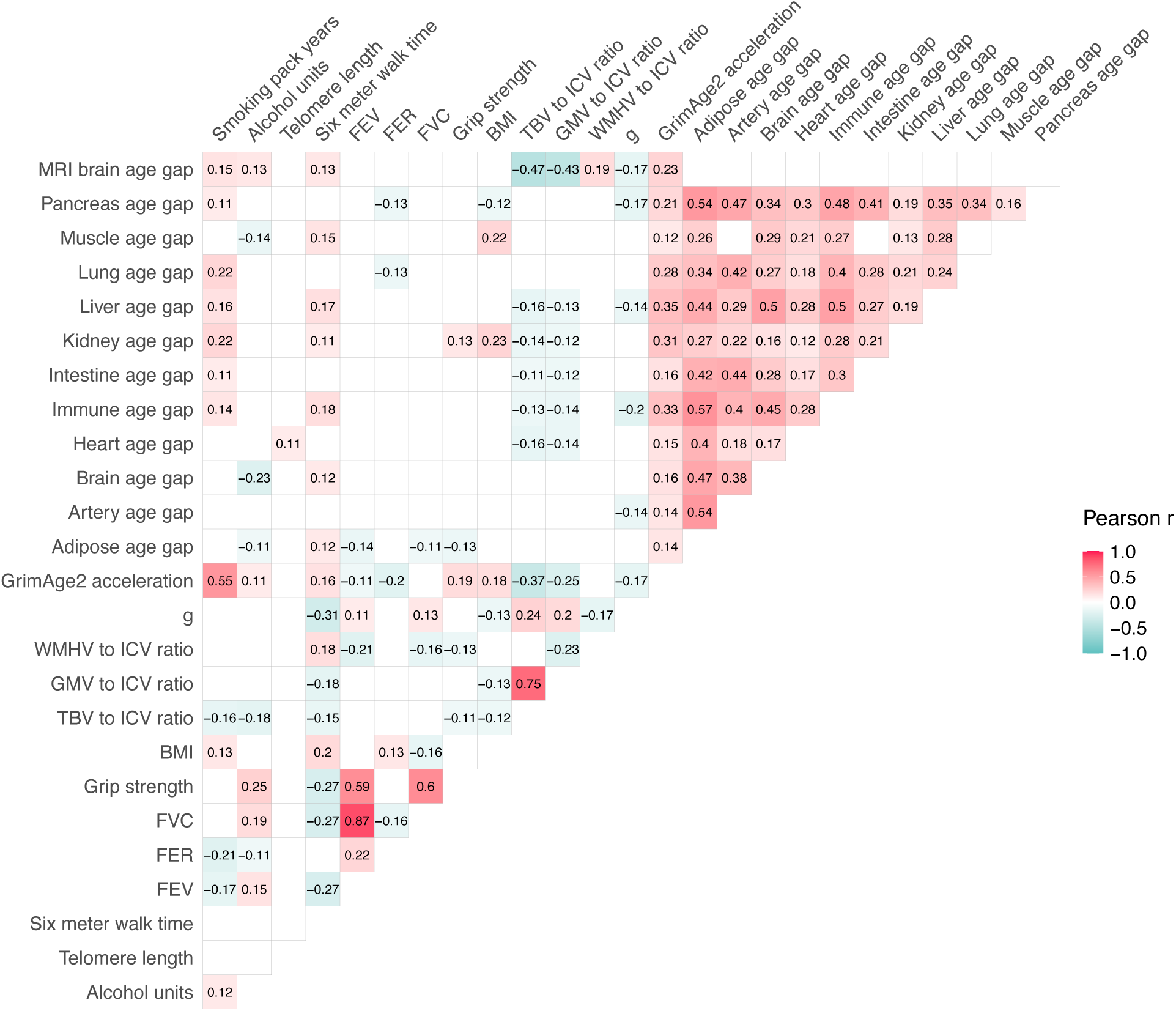
Pairwise Pearson correlations among ageing biomarkers and modifiable risk factors in LBC1936 wave 2 participants. Most biomarkers are weakly (|r| ≤ 0.4) to moderately (0.4 < |r| ≤ 0.7) correlated, with strong correlations (|r| > 0.7) observed only between forced expiratory ratio (FER) and forced expiratory volume (FEV), and between grey matter volume (GMV) and total brain volume (TBV). Red squares indicate positive correlations, and green squares indicate negative correlations. Only significant associations are displayed (P_FDR_ < 0.05).

**SUPPLEMENTARY FIGURE 3.**
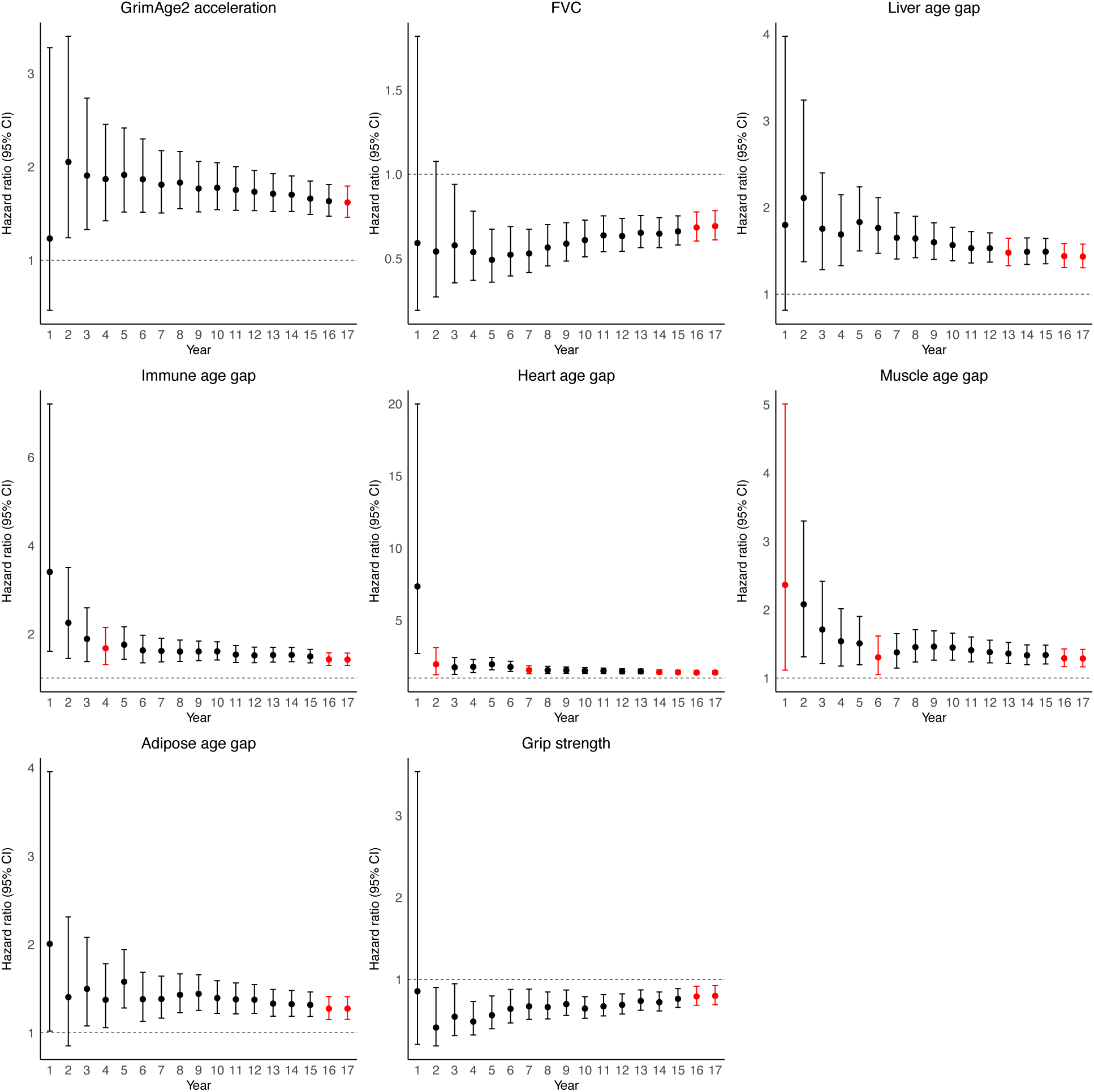
Change in hazard ratios of ageing biomarkers for all-cause mortality over up to 17 years of follow-up in the LBC1936. Cox proportional hazards regression models were used to assess associations between individual ageing biomarkers measured at wave 2 and all-cause mortality (n = 326–444 deaths in 656–861 individuals, varying by biomarker). Models were fitted annually for up to 17 years of follow-up and adjusted for chronological age and sex. Forest plots are shown only for ageing biomarkers that violated the proportional hazards assumption (Schoenfeld residuals P < 0.05), with the year(s) of violation highlighted in red. Circles represent estimated hazard ratios, with error bars indicating 95% confidence intervals. The horizontal dotted line denotes a hazard ratio of 1.

**SUPPLEMENTARY FIGURE 4.**
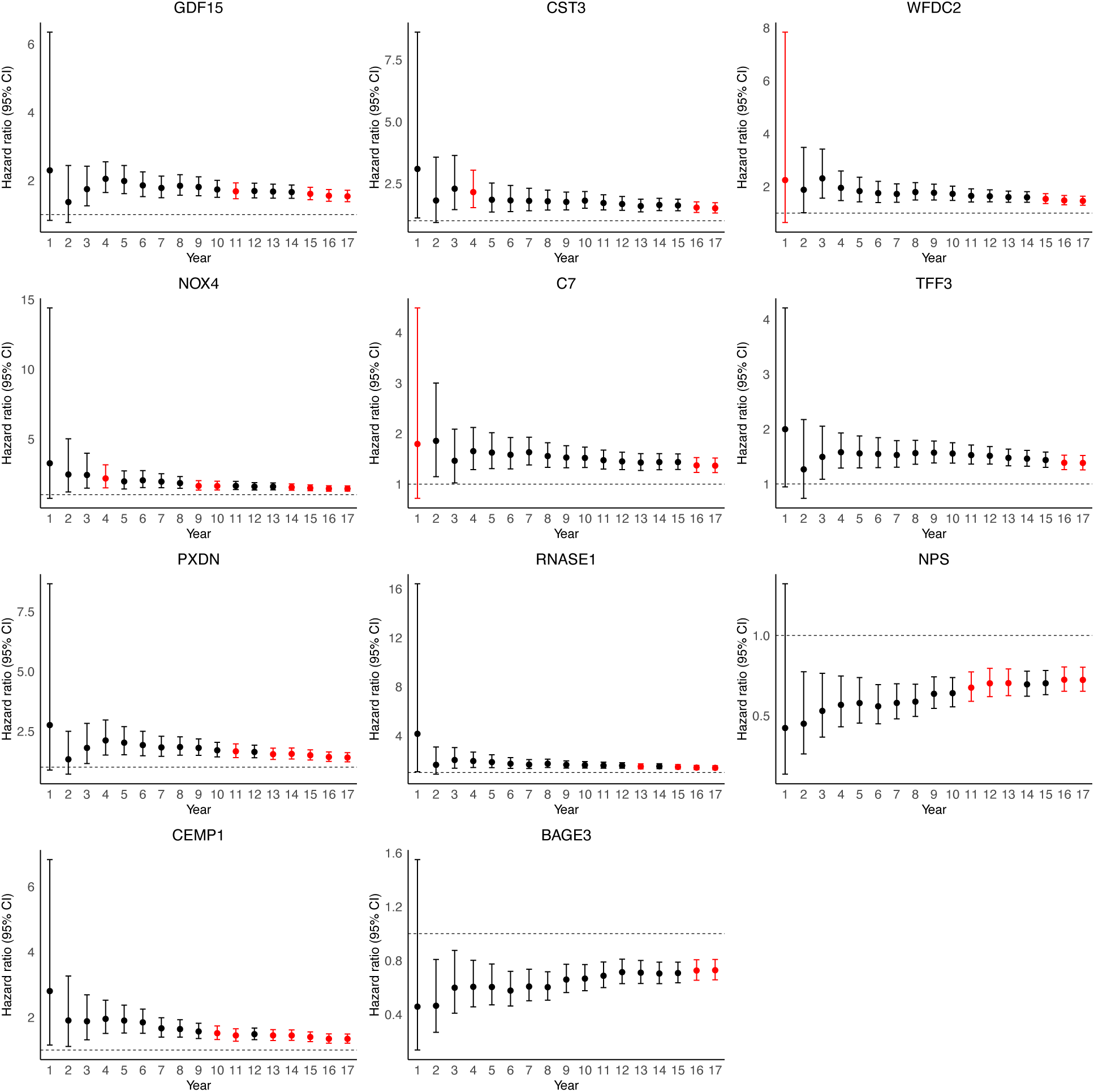
Change in hazard ratios of plasma proteins for all-cause mortality over up to 17 years of follow-up in the LBC1936. Cox proportional hazards regression models were used to assess associations between individual SomaScan 11K plasma proteins measured at wave 2 and all-cause mortality (n = 358 deaths among 694 individuals). Models were run annually for up to 17 years of follow-up and adjusted for chronological age, sex, smoking, alcohol consumption, body mass index (BMI), and estimated glomerular filtration rate (eGFR). Forest plots show 11 of the top 20 proteins with the largest hazard ratios that violated the proportional hazards assumption (Schoenfeld residuals P < 0.05), with the year(s) of violation highlighted in red. Circles represent estimated hazard ratios, and error bars indicate 95% confidence intervals. The horizontal dotted line denotes a hazard ratio of 1.

## Notes

### Author Declarations

Ethical approval for the Lothian Birth Cohort 1936 (LBC1936) study was obtained from the Scotland A Research Ethics Committee, National Health Service (NHS) Research Ethics Service, United Kingdom (reference: 07/MRE00/58). Ethical approval was granted for the collection and analysis of the cohort data. All participants provided written informed consent prior to participation.

