## Supplementary figures and images for "Multimodal Ageing Biomarkers and Plasma Proteomic Signatures Associated with All-Cause Mortality"

### Supplementary Figure 1

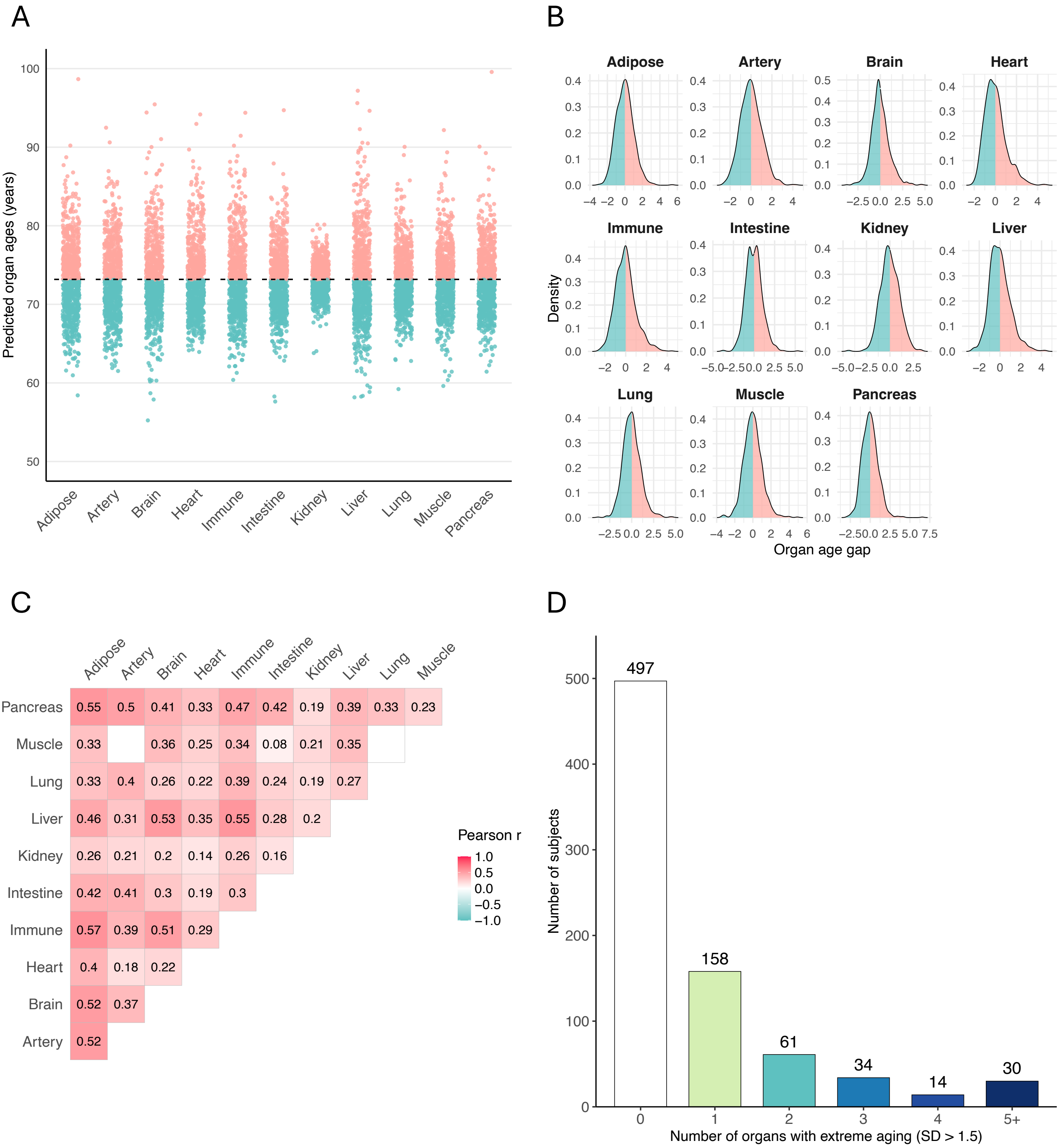

### Supplementary Figure 2

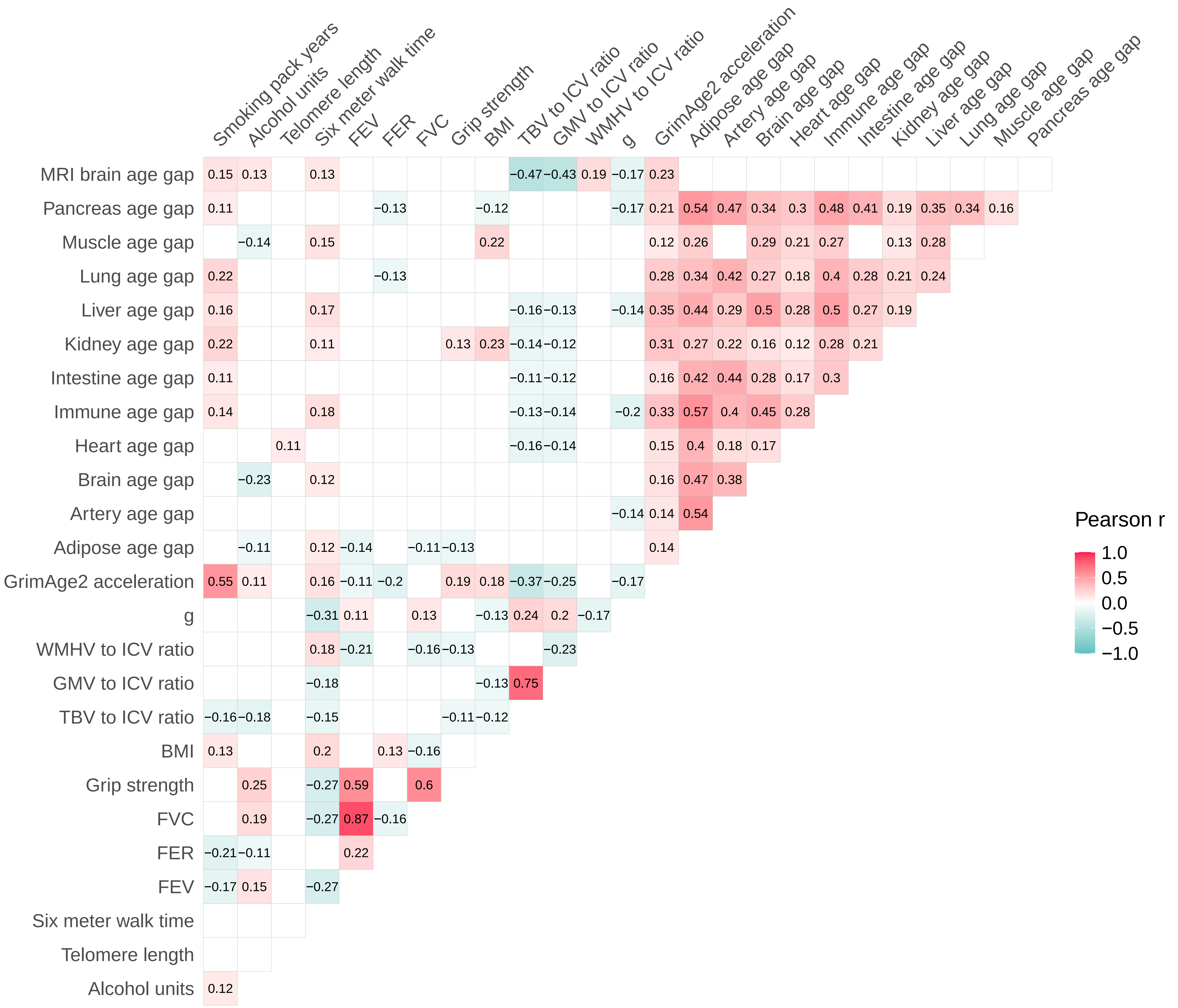

### Supplementary Figure 3

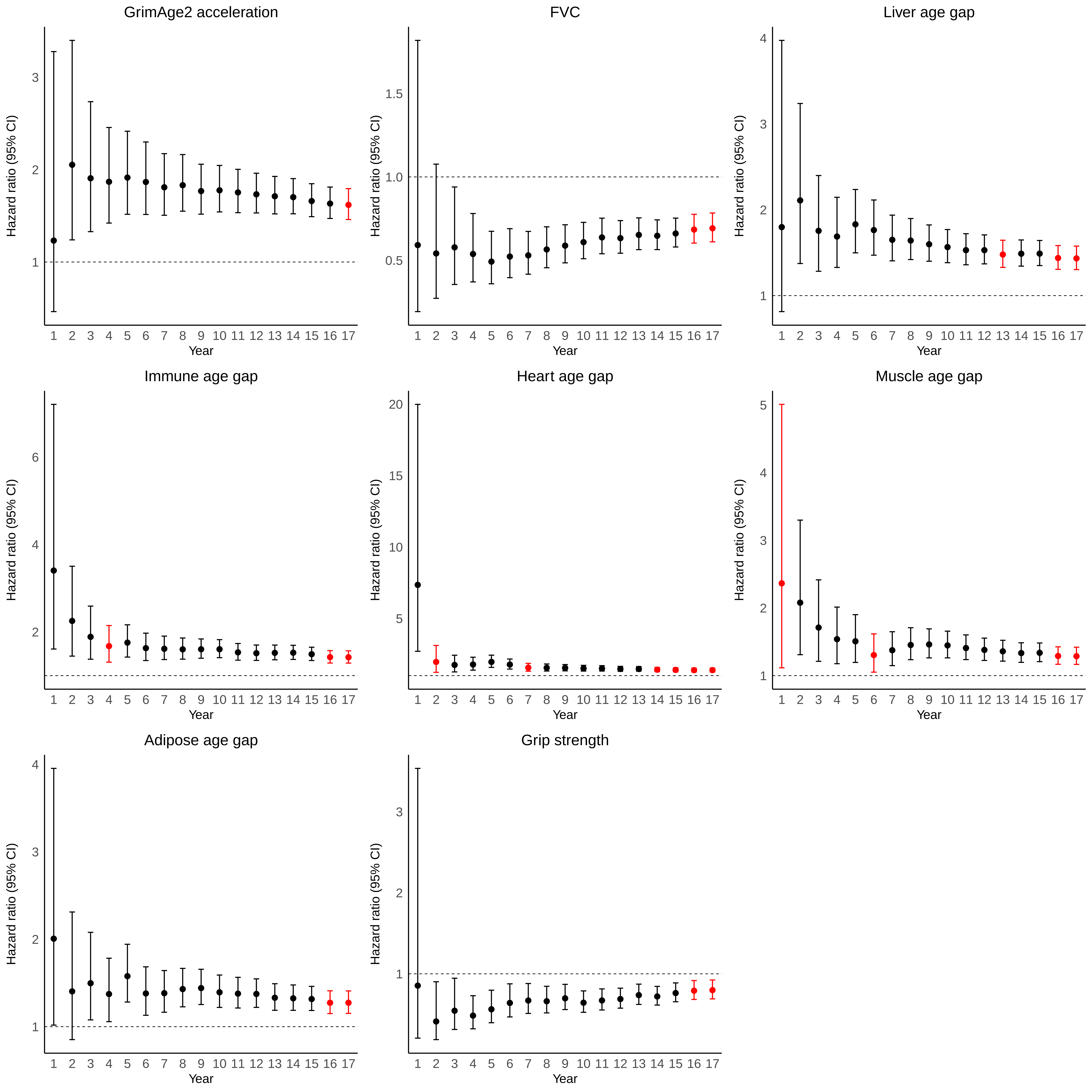

### Supplementary Figure 4

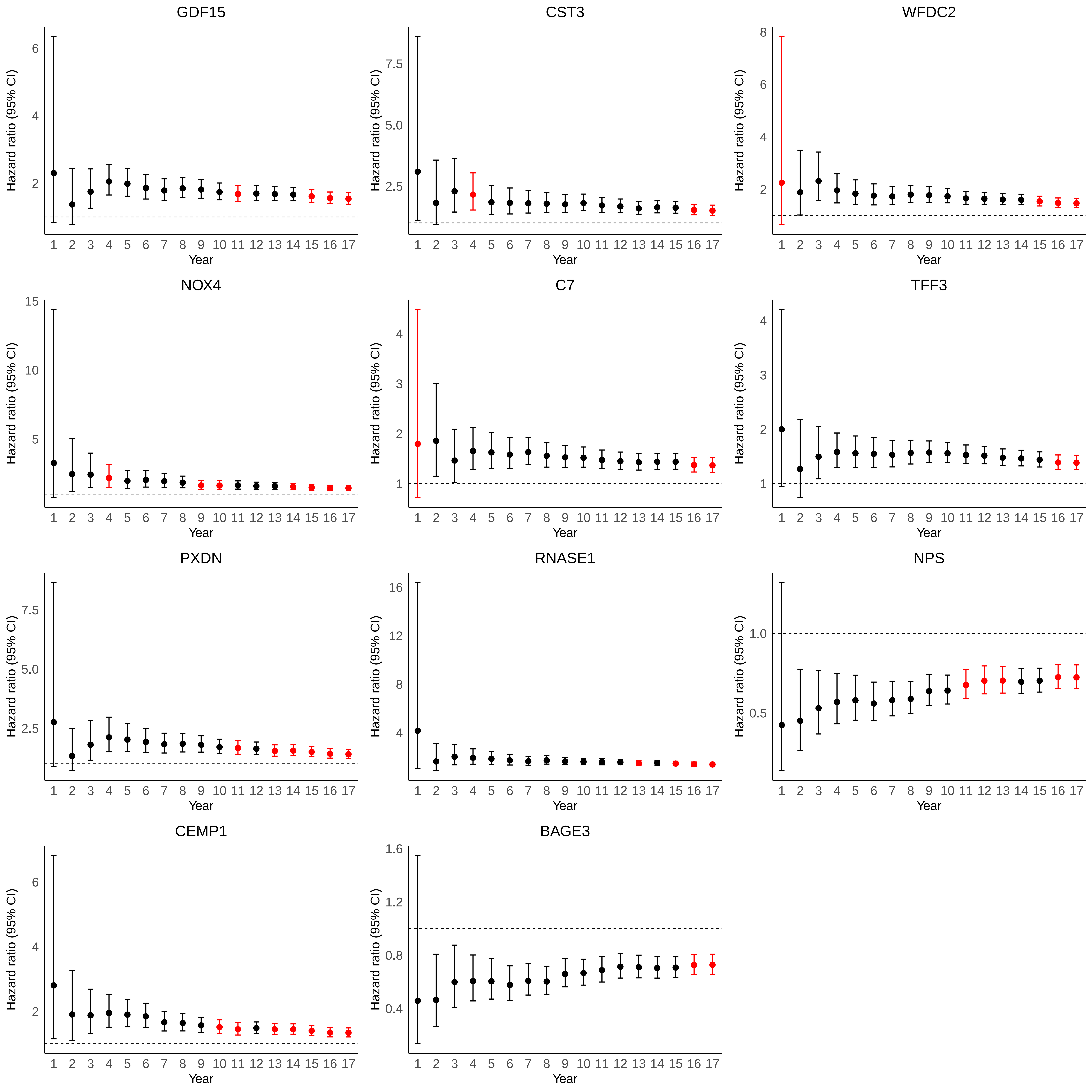
